# Researcher perspectives on the value and impact of population-based cohort studies

**DOI:** 10.64898/2026.04.06.26349895

**Authors:** Meredith O’Connor, Elodie O’Connor, Elizabeth K. Hughes, David Bann, Ken Knight, Evangeline Tabor, Charis Bridger Staatz, Sarah Gray, David P Burgner, Craig A. Olsson

## Abstract

**Background:** Population–based cohort studies are increasingly expected to demonstrate benefits for public health and wider society. However, there is limited systematic evidence on what such impact entails or how it is generated and sustained. To address this gap, we examined researcher perspectives on the impact of cohort studies.

**Methods:** We conducted, to our knowledge, the first quantitative study of researcher views on cohort impact, recruiting active cohort researchers through national and international networks between August and December 2025. The anonymous cross–sectional survey captured researcher characteristics, perceived contributions, impact processes, challenges, and open–ended reflections.

**Results:** A total of 163 cohort researchers participated, primarily from Australia (42%) and the UK (23%). Participants perceived their work as informing a wide range of societal issues and reported investing an average of 24% of their work time in impact–related activities. While most respondents (73%) believed their research leads to tangible policy or practice change, two–thirds indicated that impact is rarely or never demonstrable shortly after study completion (67%) and seldom attributable to a single study (67%). Key concerns included pressure to overstate contributions (80%), perceived disadvantages for cohort studies in impact assessments (78%), and inadequate skills or resources to achieve impact (65%).

**Conclusions:** Cohort researchers perceive their work as generating broad societal contributions and invest substantial effort in supporting impact. However, they face systemic challenges in both achieving and demonstrating impact. These findings highlight the need for impact frameworks that better capture complexity, long–term influence, and cumulative contributions, while mitigating unintended consequences.

**Key Messages:**

- Researchers perceive population–based cohort studies as generating broad societal value and contributing evidence relevant to complex societal challenges.
- Cohort impact is described as unfolding over long timescales and rarely attributable to single studies; features that sit uneasily within short–term, attribution–focused impact models.
- Current impact frameworks appear poorly aligned with long–term research investments and may create unintended pressures to overstate research contributions.

---

Population-based prospective cohort studies tracking large groups of individuals over time are foundational to multiple disciplines, including epidemiology.^1^ They offer unique opportunities to identify long–term influences on health outcomes that cannot be feasibly or ethically examined using randomised trials, cross–sectional designs, or administrative data alone.^2, 3^ Alongside their scientific value, cohort studies are increasingly expected to demonstrate their contribution to addressing the many societal challenges affecting population health and wellbeing.^4, 5^

Assessment of these contributions, however, tends to privilege what is easily measured and directly attributable, favouring short–term, linear, and discrete impact outcomes.^6^ In contrast, cohort studies are designed for long–term inquiry, functioning as enduring research infrastructure that generate systemic and often unforeseen contributions^7, 8^ that are poorly captured by traditional metrics.^9^ The development of life–course theory, now foundational to countless health and social policies, illustrates this form of impact through sustained influence within dynamic policy systems.^8^

Understanding cohort impact also requires attention to the processes through which it is generated.^10, 11^ Activities such as media engagement, cross–sector collaboration, and parliamentary outreach are often assumed to facilitate translation into action.^11, 12^ Yet without evidence on the extent to which researchers use these approaches, particularly in informal or less visible ways, it is difficult to assess their relative effectiveness. In addition, potential unintended consequences, such as sensationalism or excessive researcher burden, may be overlooked.^13, 14^

Researchers are not passive conduits of evidence but active agents who invest substantial time and effort in generating impact,^10–12, 15^ informed by their disciplinary and research contexts.^16^ Recent evidence shows that public health researchers increasingly discuss policy implications in their cohort publications,^14^ consistent with qualitative insights that most cohort researchers value and actively pursue impact.^17^ These emerging insights highlight both the feasibility and importance of developing an impact evidence base that is grounded in the realities of cohort research.

## The current study

Despite rising expectations for cohort studies to demonstrate impact, evidence on the nature of impact in this context or how such impact is generated and sustained remains limited. To address this gap, we conducted, to our knowledge, the first survey of cohort researchers to quantify their views and experiences. The survey aimed to address three questions:

1. Types of impact: What contributions do cohort researchers perceive in their work?
2. Impact processes: How does impact typically unfold, and what strategies do cohort researchers use in their efforts to generate and maximise impact?
3. Challenges and opportunities: What challenges and opportunities do cohort researchers perceive in relation to impact?

## Methods

### Data source

Data were collected through an anonymous cross–sectional online survey of cohort researchers (Supplementary 1), administered via REDCap.^18^ The study was approved as negligible risk by the Royal Children’s Hospital (Melbourne, Australia) Human Research Ethics Committee.

To maximise reach, we disseminated survey invitations through emails, newsletters, LinkedIn, and publicly available contact lists from multiple cohort collaborative networks with overlapping subscriber bases. Participants were also encouraged to share the invitation within their own networks using snowball sampling. This approach precluded a formal response rate but enabled broad participation across a highly distributed field.

Any researcher actively engaged with cohorts in the past 12 months was eligible to participate. Eligibility was confirmed in three ways:

1. The survey invitation outlined eligibility criteria and provided broad definitions of a cohort and cohort researcher; see Supplement 1.
2. A mandatory screening question at the beginning of the survey asked about respondents’ role/s in cohort research over the past 12 months; seven respondents were excluded at this stage because they had not undertaken a relevant role in that period.
3. Reported cohort characteristics were reviewed to confirm alignment with key features of cohort studies (e.g. coverage of different life stages).

### Measures

We adapted items from the few existing instruments examining researcher views on impact in other fields of research,^16, 19, 20^ and created new items where no suitable measures existed. Items were piloted and refined with a small group of cohort researchers. The final survey (Supplementary File 1; summarised in Table 1) included: researcher and cohort characteristics; perceived contributions of cohort research; views on the impact process and impact strategies they employ; perceived challenges and opportunities; and open-ended reflections.

**Table 1.** Summary of survey content and approach.

| Topic | Question | Items | Provenance | Response scale |
| --- | --- | --- | --- | --- |
| **Researcher and primary cohort characteristics** |  |  |  |  |
| Researcher role | Over the past 12 months, which of the following best describes your involvement with cohort research? Tick all that apply. By "cohort", we are referring to population-based longitudinal studies that collect data about a group of individuals over time. | Senior leadership as a cohort director, chief or associate investigator, or similar; Project coordination and operations; Lived experience/consumer advisor; Analysis and research using cohort data; Other active role/s in cohort research; No involvement in cohort research in the past 12 months [discontinues survey] | Study derived | Check all that apply |
| Years in cohort research | How long have you been working in cohort research? |  | Adapted from <sup>16</sup> | Round to the nearest whole year |
| Primary country in which working | What country do you primarily work in? |  | Adapted from <sup>16</sup> | Select country name |
| Discipline | Which of the following best describe the discipline of your cohort research? | Biostatistics and data science; Demography; Economics; Education; Epidemiology and public health; Genetics and molecular biology; Medicine and biomedical sciences; Psychology; Sociology; Other | Adapted from <sup>20</sup> and <sup>16</sup> | Tick all that apply |
| Concurrent appointments | Do you hold any concurrent appointments outside of the research/academic sector? |  | Study derived | No, I do not hold any concurrent appointments outside the research/academic sector; Clinical (e.g., medical doctor, psychologist, allied health professional); Government (e.g., policy analyst, public sector strategist); Corporate (e.g., consultancy, pharmaceuticals industry); Nonprofit (e.g., social impact initiatives, philanthropic organisation); Other role outside of the research / academic sector |
| Life periods captured | What life periods does the cohort capture? |  | Study derived | Childhood (pre-conception to 10 years); Adolescence (11-25 years); Young adulthood (26-39 years); |
|  |  |  |  | Middle adulthood (40-59 years);<br>Later adulthood (60-79); Senior adulthood (80+) |
| Sample size of cohort at recruitment | What is the approximate sample size (number of participants recruited at baseline) being followed within this cohort study? |  | Study derived | 499 or fewer participants; 500-999 participants; 1000-4,999 participants; 5000-9,999 participants; 10,000-19,999 participants; More than 20,000 participants |
| Primary country in which cohort is based | Where is the cohort primarily based? |  | Study derived | Select country name |
| **Impact types** |  |  |  |  |
| Domains of impact | To what extent does your cohort research contribute across the following domains? We are interested in your views on the current focus and objectives of your cohort research, rather than measurable progress. | Research (e.g., pursuit of new knowledge, developing new research methods or approaches, shaping the research agenda in your field, or training of junior researchers); Policy (e.g., driving development of new policy, raising awareness and mobilising support for an issue among policymakers, or introducing new ideas, language, or concepts into policy discussions); Services (e.g., change to treatment or targeting of intervention, informing data and reporting in health, education, social, or other service systems, improving the cost-effectiveness of services); Societal (e.g., shaping public understanding and awareness of issues, influencing societal values and culture, and improving the public's ability to access and use information that benefits them); Other | Study derived with items aligned to health impact frameworks<sup>21, 38</sup>, informed by format of similar item set in <sup>19</sup> | Not at all; Slightly; Moderately; Strongly; Not sure |
| Societal issues addressed | To what extent does your cohort research focus on the following | Health and wellbeing (e.g. mental and physical health, substance use); Family | Study derived item informed by format of | Not at all; Slightly; Moderately; Strongly; Not sure |
|  | societal issues? We are interested in your views on the current focus and objectives of your cohort research, rather than measurable progress in tackling these large-scale social issues. | relationships (e.g. parenting, family functioning); Education (e.g. curriculum, bullying, attainment); Workplace wellbeing (e.g. stress, career pathways); Community and social networks (e.g. social cohesion, peer support); Crime and justice (e.g., delinquency, victimisation, contact with criminal justice system); Economic factors (e.g. cost of living, housing availability); Social equity (e.g. discrimination, social disadvantage); Digital and technology (e.g. online safety, cybersecurity); Historical impacts (e.g. intergenerational trauma, population aging); Other | similar item set in <sup>16</sup>. Relevant items were developed based on issues of greatest importance to young people<sup>22</sup> and adult Australians<sup>23</sup>, global risks in the World Economic Forum review,<sup>24</sup> and models underpinning cohort studies.<sup>25, 26</sup> |  |
| **Impact processes** |  |  |  |  |
| Views on broader impact processes | Thinking about the impact of your cohort research, how often do you observe the following? We are interested in your perceptions of how impact typically unfolds, based on your experience in cohort research. | The research leads to specific, tangible changes in policy or practice; It's possible to trace a clear, causal link between cohort research and its impact; Impact can be demonstrated shortly after the research is conducted; Impact arises from a single piece of research (as opposed to a body of work) | Study derived | Never; Rarely; Sometimes; Almost always; Not sure |
| Strategies to generate impact | What strategies does your cohort research program employ to try to achieve impact? We do not expect all cohorts to employ all strategies, nor do we assume that these strategies necessarily translate into measurable impact. | Strategic planning (e.g., co-defining impact goals with stakeholders; planning and embedding impact evaluation); Academic dissemination (e.g., peer-reviewed articles; conferences or symposia; segments in institutional newsletters); Policy reports (e.g., producing summaries of evidence in relation to policy, contributing submissions to government inquiries); Community forums (e.g., accessible talks, workshops or training sessions for community groups); Traditional media (e.g., sharing evidence through broadcast or print via major news outlets); Digital and social media (e.g., podcasts, LinkedIn posts, Twitter/X threads, short-form videos); Cross-sector partnerships (e.g., undertaking research with industry or NGO partners); Parliamentary engagement (e.g., roundtables that connect researchers and policymakers); Other | Question stem adapted from <sup>19</sup> and <sup>16</sup>, items adapted from <sup>11</sup> and <sup>27</sup> | Never; Rarely; Sometimes; Almost always; Not sure |
| Time investment | What time investment does impact require? The activities above can involve time intensive tasks such as meetings or emails with stakeholders, development of translational outputs, and strategising about impact. | In a typical week, what proportion of your total work time do you spend on these activities? In an ideal week, what percentage of your work time would you like to spend on these activities? | Study derived | Slider selecting response from 0% to 100% |
| **Challenges and opportunities** |  |  |  |  |
| Views and attitudes about impact in the context of cohort research | To what extent do you agree with the following statements about how research impact is currently approached? We are interested in your views on the current research environment, which includes research policy, funders, journals, and research organisations. | Current impact approaches align well with the goals and motivations of cohort research; Existing impact frameworks capture and value the types of societal impact that cohort studies typically generate; Pressure exists for cohort researchers to overstate claims about impact; Cohort studies are disadvantaged compared to other study designs by current impact evaluation approaches; Skills, resources, and support are available to cohort teams to plan, enable, and monitor impact | Informed by similar item sets in <sup>16, 20</sup> | Strongly disagree; Somewhat disagree; Somewhat agree; Strongly agree; Not sure |
| **Open-ended reflections** |  |  |  |  |
| Remaining reflections | Would you like to offer any other reflections regarding impact in the context of cohort studies? For example, we are interested to hear about other barriers you face, whether you feel adequately resourced in this area, how you are approaching specific areas like monitoring your impact, or specific examples of impact that you may wish to share. |  | Study derived | Free text box |

The survey deliberately focused on eliciting researchers’ subjective perceptions, to align with our research questions on perceived impact, practices, and attitudes. Complete detachment of participants from the topic was not possible, but to minimise social desirability respondents were repeatedly reminded that the study sought their personal views and that all responses were anonymous. Because cohort researchers occupy diverse roles, respondents were instructed to answer with reference to their cohort research program as they define it, ensuring relevance for participants ranging from senior investigators to secondary data users.

*Researcher and cohort characteristics:* We collected demographic data, including role type, years working in cohort research, primary discipline, concurrent appointments outside academia, and country of residence. Respondents also reported features of the cohort that was the primary focus of their active role(s) over the past 12 months, including life periods covered, sample size, and geographic context.

*Types of impact:* Perceived impact domains (e.g. policy sector, service sector) were assessed using a 4-point scale (1=Not at all to 4=Strongly), based on existing health impact frameworks^21^ and a question stem from prior research.^19^ Additional items asked respondents to rate the extent to which their cohort research addressed specific societal issues (e.g. health and wellbeing, social equity, education), informed by prior instruments,^16^ community priorities,^22, 23^ global risks,^24^ and cohort theoretical models.^25, 26^

*Impact processes:* New items measured researcher views on characteristics of the impact process (e.g. Impact can be demonstrated shortly after the research is conducted; 1=Never to 4=Almost always). To examine strategies used to generate impact, we adapted and extended an item set from prior work,^16^ focusing on activities within researcher control and highlighted in existing literature (e.g. strategic planning, publications for diverse audiences).^11, 27^ Respondents indicated frequency of use on a 4-point scale (1=Never to 4=Almost always). Participants were also asked to estimate the proportion of work time dedicated to these activities in a typical week and an ideal week.

*Challenges and opportunities:* Items adapted from previous surveys^16, 20^ assessed agreement with potential challenges and opportunities specific to cohort research (e.g. Existing impact frameworks capture and value the types of societal impact that cohort studies typically generate) using a 4-point scale (1=Strongly disagree to 4=Strongly agree).

*Open ended questions:* Participants were invited to share additional reflections in a free–text format.

### Statistical analysis

We examined missingness across all variables (Supplementary 2, Table 1) and found low levels for participant demographics (range=0-3%; M=1%), perceived impact (range=1-6%; M=3%), and impact processes (range=1-6%; M=4%). Missingness was higher for items on challenges and opportunities (range=9-15%; M=13%), primarily due to “not sure” responses, which were recoded as missing. All available data from N=163 eligible respondents were analysed.

Sample characteristics and cohort features were summarised using counts and percentages, and main survey variables as percentages with 95% Confidence Intervals. To protect anonymity, category counts with fewer than five respondents were collapsed into “other.” Percentages were visualised in figures, with full estimates reported in supplementary tables (Supplementary 2).

Qualitative responses were analysed thematically to supplement quantitative findings. The lead author reviewed all responses to develop an initial coding framework, with blind–coding by author EO’C for validation. Discrepancies were resolved through discussion. To ensure anonymity and relevance, only themes supported by more than five participants are reported.

## Results

### Researcher characteristics

A total of 163 respondents contributed data (56% analysis/research; 38% senior leadership; 30% operations; Table 2). Respondents were primarily based in Australia (42%) and the UK (23%), representing 2,061 cumulative years of cohort research experience. Most (80%) had disciplinary backgrounds in epidemiology or public health, and around one–fifth held concurrent non–academic appointments, such as clinical practice (9%).

**Table 2.** Characteristics of participating cohort researchers and the cohorts they primarily work with (N=163).

| Characteristics | N (%) |
| --- | --- |
| **Researcher characteristics** |  |
| **Role type<sup>^</sup>** |  |
| Senior leadership and management as a cohort director, chief or associate investigator, scientific advisor, or similar | 62 (38.0%) |
| Analysis and research using cohort data | 91 (55.8%) |
| Project coordination and operations | 49 (30.1%) |
| Other active role/s in cohort research<sup>a</sup> | 14 (8.6%) |
| **Years in cohort research** |  |
| 0-5 | 54 (33.1%) |
| 6-10 | 32 (19.6%) |
| 11-15 | 31 (19.0%) |
| 16-20 | 15 (9.2%) |
| 20+ | 31 (19.0%) |
| **Primary country in which working** |  |
| Australia | 68 (41.7%) |
| United Kingdom | 38 (23.3%) |
| France | 18 (11.0%) |
| Canada | 13 (8.0%) |
| Other | 26 (16.0%) |
| **Field of Research<sup>^</sup>** |  |
| Epidemiology and public health | 131 (80.4%) |
| Psychology | 55 (33.7%) |
| Biostatistics and data science | 35 (21.5%) |
| Medicine and biomedical sciences | 30 (18.4%) |
| Education | 26 (16.0%) |
| Sociology | 19 (11.7%) |
| Genetics and molecular biology | 16 (9.8%) |
| Demography | 11 (6.7%) |
| Economics | 7 (4.3%) |
| Other | 7 (4.3%) |
| **Concurrent appointments<sup>^</sup>** |  |
| No concurrent appointments outside academia/research | 126 (77.3%) |
| Clinical (e.g., medical doctor, psychologist, allied health professional) | 15 (9.2%) |
| Nonprofit (e.g., social impact initiatives, philanthropic organisation) | 8 (4.9%) |
| Government (e.g., policy analyst, public sector strategist) | 7 (4.3%) |
| Other<sup>b</sup> | 5 (3.1%) |
| **Cohort the researcher primarily works with** |  |
| **Primary country in which cohort is based** |  |
| Australia | 66 (41.5%) |
| United Kingdom | 35 (22.0%) |
| France | 16 (10.1%) |
| Canada | 13 (8.2%) |
| Other | 29 (18.2%) |
| **Life periods captured<sup>^</sup>** |  |
| Childhood (pre-conception to 10 years) | 99 (60.7%) |
| Adolescence (11-25 years) | 96 (58.9%) |
| Young adulthood (26-39 years) | 73 (44.8%) |
| Middle adulthood (40-59 years) | 65 (39.9%) |
| Later adulthood (60-79) | 54 (33.1%) |
| Senior adulthood (80+) | 34 (20.9%) |
| **Sample size of cohort at recruitment** |  |
| <1000 participants at baseline | 16 (9.9%) |
| 1000-4,999 participants | 46 (28.4%) |
| 5000-9,999 participants | 32 (19.8%) |
| 10,000-19,999 participants | 34 (21.0%) |
| More than 20,000 participants | 34 (21.0%) |

^Sum to greater than 100% as multiple responses could be selected. <sup>a</sup>Other includes “Lived experience/consumer advisor”, combined due to cell size<5. <sup>a</sup>Other includes “Corporate (e.g., consultancy, pharmaceutical industry)”, combined due to cell size<5.

### Cohort characteristics

Participants’ work focused on cohorts predominantly based in Australia (42%) and the UK (22%). These cohorts covered diverse life stages from childhood (61%) to senior adulthood (21%), and ranged in size from fewer than 1,000 participants (10%) to more than 20,000 participants (21%).

### Types of impact

When asked about the extent to which their cohort research contributed to four domains, the research domain was most frequently identified as a major area of contribution (moderately: 11%, CI=7-17; strongly: 84%, CI=77-89; Figure 1, Supplementary 2 Table 2). Beyond research, contributions were most frequently reported in the policy domain (moderately: 46%, CI=39-54; strongly: 31%, CI=24-39), followed by the general public (moderately: 41%, CI=33-48; strongly: 27%, CI=20-34), and service systems (moderately: 35%, CI=28-43; strongly: 19%, CI=14-26).

**Figure 1.**
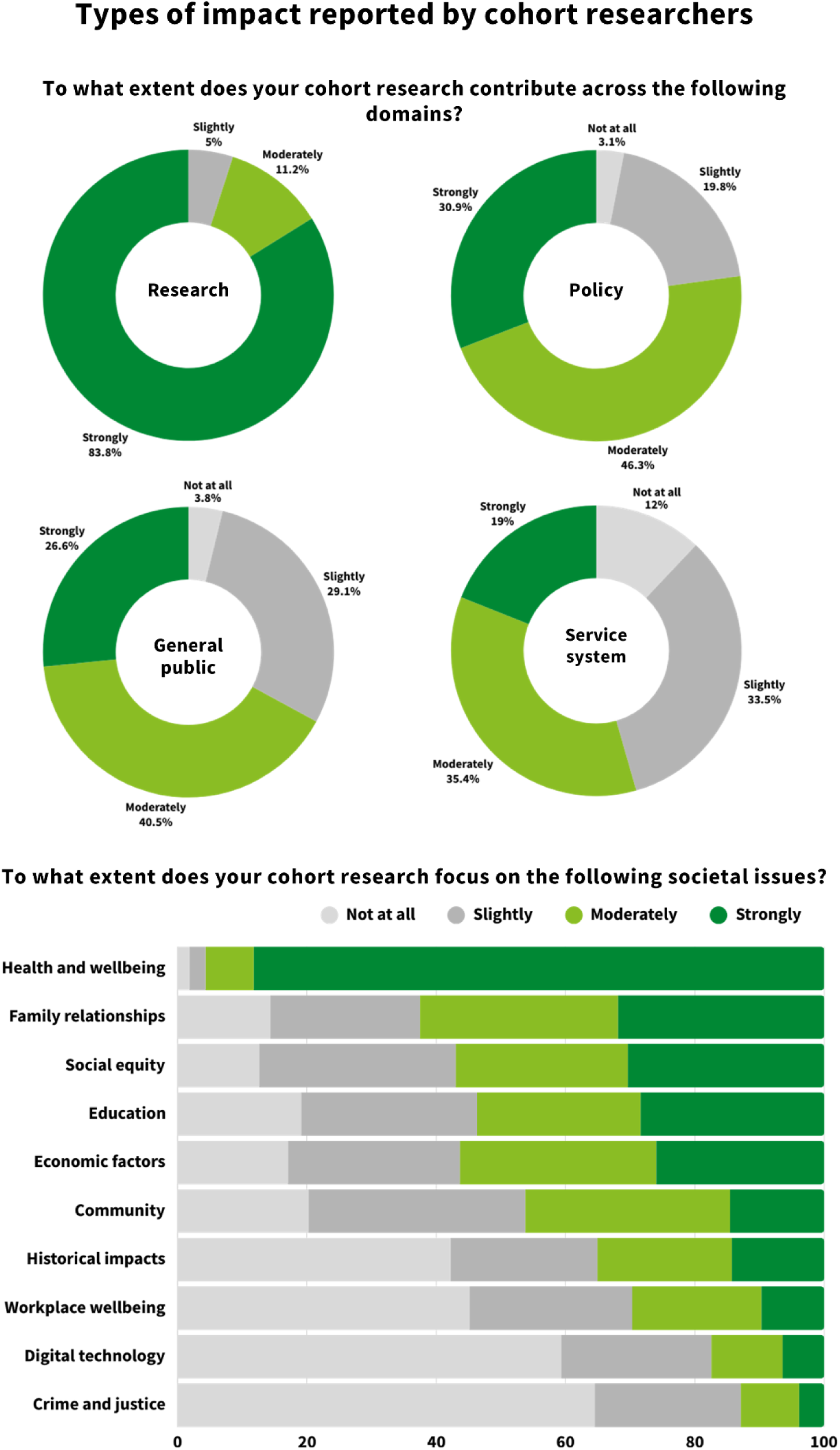
Types of impact reported by cohort researchers based on their subjective views of the current focus and objectives of their cohort research.

When considering specific societal issues, health and wellbeing was the strongest area of perceived contribution (moderately: 7%, CI=4-13; strongly: 88%, CI=82-92). This was followed by family relationships (moderately: 31%, CI=24-38; strongly: 32%, CI=25-40), social equity (moderately: 27%, CI=20-34; strongly: 30%, CI=24-38), and education (moderately: 25%, CI=19-33; strongly: 28%, CI=22-36). Free-text responses identified additional areas of contribution, including climate change, the built environment, and cultural identity.

### Impact processes

Three-quarters of participating researchers believed their work achieves tangible policy or practice change (sometimes: 65%, CI=57-72; almost always: 8%, CI=4-13; Figure 2, Supplementary 2 Table 3), and two-thirds reported that it was possible to trace a clear causal link between research and impact (sometimes: 56%, CI=48-63; almost always: 9%, CI=6-15). However, most disagreed that impact is demonstrable shortly after completion (rarely: 58%, CI=50-66; never: 9%, CI=5-15) or that it typically arises from a single study (rarely: 50%, CI=42-58; never: 17%, CI=12-24).

**Figure 2.**
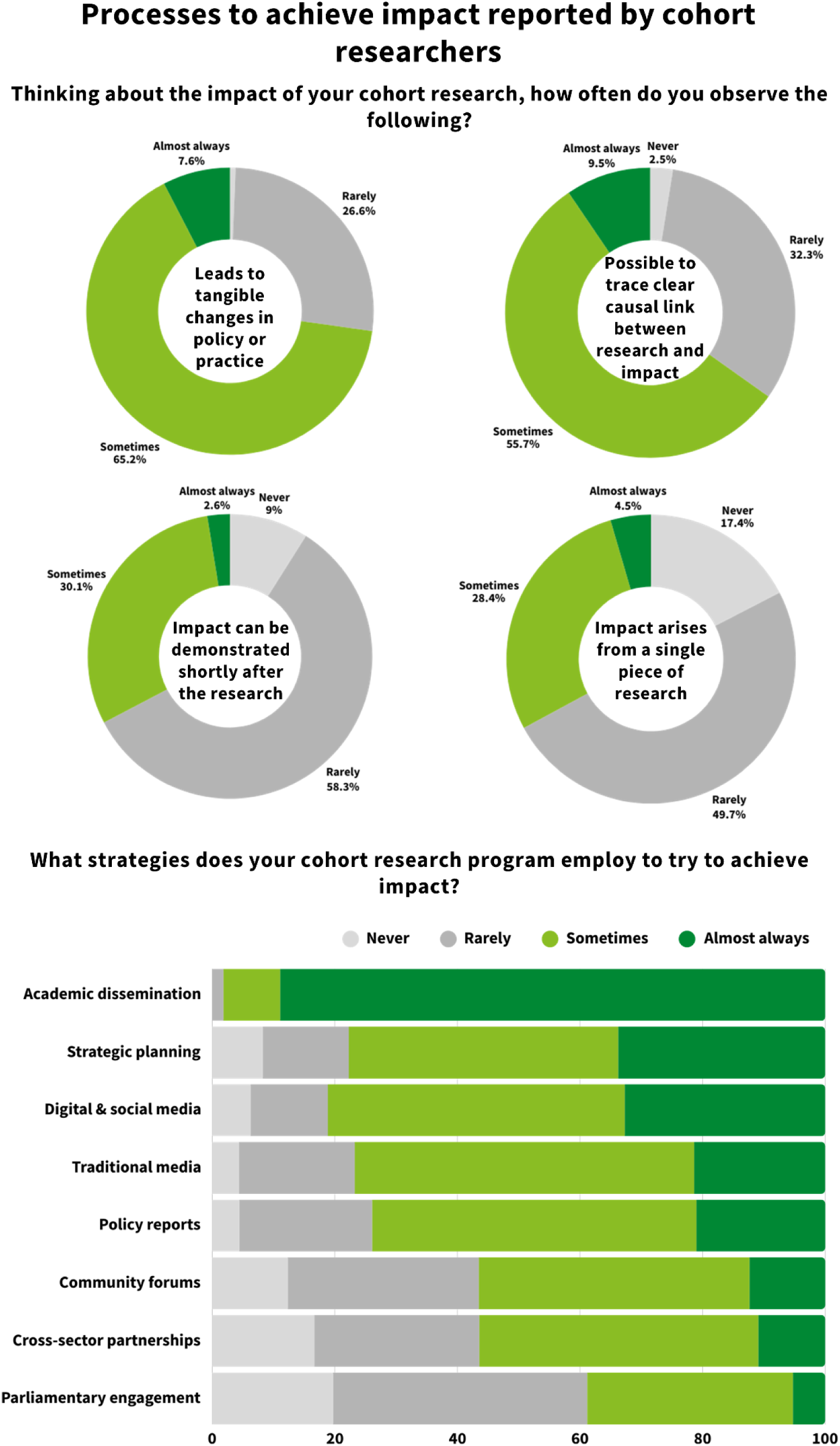
Processes to achieve impact reported by cohort researchers, based on their subjective viewpoint.

**Table 3.** Key themes derived from free text responses offering additional thoughts and reflections (N=43).

| Theme<sup>^</sup> | Description of theme | N (%) | Example |
| --- | --- | --- | --- |
| Funding system tensions and suggestions | Reflections on the broader funding system highlighted the tension between short-term funding cycles and the long-term nature of cohort studies, which was seen as affecting all aspects of cohort management and research, including impact. Funder expectations were often perceived as misaligned with cohort realities, emphasizing short-term outcomes over long-term influence, overlooking the importance of discovery research, and favouring direct, RCT-style results. Suggested solutions included introducing long-term monitoring of impact outcomes for funded projects. | N=22, 52% | "...The current funding structure stands as a formidable barrier, preventing most large cohorts from ever truly realising their intended potential. Without meaningful change in the cohort funding sector, we risk perpetually spinning our wheels trapped in the same cycle, devoid of genuine progress, measurable achievements, and transformative outcomes." |
| Specific areas of resourcing and support needs | Respondents noted how funding tensions showed up in their everyday practice to drive specific areas of resource needs. Despite noting that excellent examples exist, concerns were raised that resource constraints often result in impact activities being post-hoc, rushed, and lacking strategic intent. Respondents called for greater support, including education, hands-on assistance, and clearer guidance on what an impact strategy should entail. Respondents also noted that these types of impact activities require time and resources rarely budgeted for, leaving this work unpaid and often undertaken in researchers' own time. | N=18, 43% | "...There are still a lot of rushed, post hoc activities which may not be purposeful and miss the mark. There is room for education and hands-on support on what a research impact strategy actually entails and how to build this into projects from the outset." |
| Difficulties and challenges of impact work in the cohorts context | Respondents described cohort impact work as particularly challenging. The discovery-oriented nature of cohort research, often focused on addressing complex problems, was described as involving extended impact pathways and emerging through the cumulative weight of a body of evidence. These challenges were seen as further compounded by the operating conditions of cohort studies, including short policy decision cycles with limited opportunities for researcher engagement. | N=11, 26% | "Impact is too often assumed to result from single studies, rather than appreciating the complexity of how knowledge gained from cohort studies typically adds to or expands on existing knowledge. It feels that this greatly disadvantages cohort studies viz a viz clinical research or clinical trials where a more tangible result (eg contribution to a clinical guideline) might be expected." |
| Evaluation and tracking of impact is a specific challenge | Tracking and evaluating impact were identified as a particular challenge. Respondents expressed a need for metrics tailored to cohort studies, capable of capturing outcomes emerging from multiple pieces of work rather than single "silver bullet" findings. They emphasized that systemic, long-term impact cannot be fully captured by current frameworks and some advocated for common approaches to monitoring impact across cohorts under a consensus framework. | N=6, 14% | "...population-based longitudinal cohort studies generate a broad spectrum of impact-from advancing research and education to uncovering intergenerational determinants of health and disease. This wide-ranging, systemic impact cannot be fully captured by current metrics or software solutions." |

<sup>^</sup>Themes are not intended to be mutually exclusive.

Researchers reported using a wide range of strategies to facilitate impact (Figure 2, Supplementary 2 Table 3). Beyond the near-universal use of academic dissemination (sometimes: 9%, CI=6-15; almost always: 89%, CI=83-93), common strategies included strategic planning (sometimes: 44%, CI=36-52; almost always: 34%, CI=27-42), digital or social media engagement (sometimes: 48%, CI=41-56; almost always: 33%, CI=26-41), and producing policy reports (sometimes: 53%, CI=45-61; almost always: 21%, CI=15-28).

On average, respondents reported spending a quarter of their weekly work time on impact-related activities (M=24%, SD=23), with substantial variation across individuals (range: 0-100%) and positive skew (Supplementary 2 Figure 1). In an ideal week, they indicated they would allocate roughly one-third of their time to these activities on average (M=33%, SD=24), again with a full range (0-100%).

### Challenges and opportunities

Participating researchers reported several challenges associated with cohort research impact (Figure 3, Supplementary 2 Table 4). Around two-thirds agreed that the current focus on impact aligns well with the goals of cohort research (somewhat: 47%, CI=38-55; strongly: 12%, CI=8-19). However, a similar proportion disagreed that existing frameworks capture the value and types of societal impact typically generated by cohort studies (somewhat: 50%, CI=41-58; strongly: 13%, CI=8-20). The majority of participants reported that pressure exists to overstate impact claims (somewhat: 50%, CI=42-58; strongly: 30%, CI=23-37), and perceived cohorts to be disadvantaged compared to other designs under current approaches (somewhat: 50%, CI=42-58; strongly: 28%, CI=21-36). Over two thirds disagreed that sufficient skills, resources, and supports were available to plan, enable, and monitor impact in cohort research (somewhat: 37%, CI=29-45; strongly: 28%, CI=21-36).

**Figure 3.**
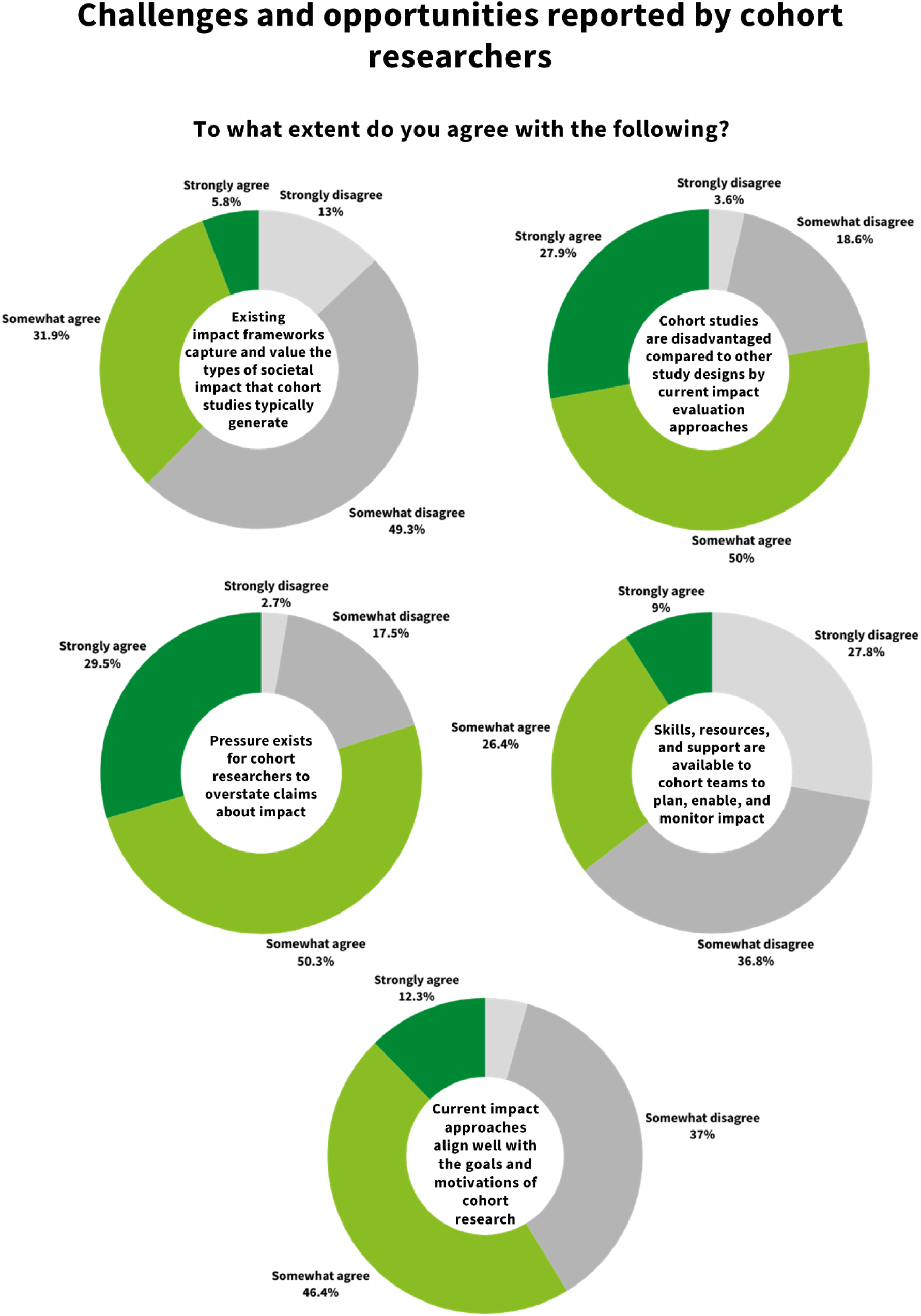
Challenges and opportunities in impact as reported by cohort researchers.

### Open ended reflections

Approximately one–quarter of respondents (N=43; 26%) provided additional reflections, with several concerns emerging. The most common theme (52%) related to tensions between short–term funding cycles and the long–term nature of cohort studies. Resource constraints were also frequently raised (43%), with respondents emphasising the need for capacity building and clearer guidance. Around one–quarter (26%) described the challenges of achieving impact in cohort studies, highlighting the inherent complexity and long time horizon. A further 14% identified evaluation of impact as a specific area requiring improved support and methodological development.

## Discussion

Expectations to demonstrate research impact are now embedded within policy and funding systems in countries such as Australia and the UK.^7, 8^ However, traditional impact metrics often fail to capture the full value of long-term cohort infrastructure.^9^ In this context, both funders^4^ and researchers^7, 8^ have called for clearer articulation of the strategic value and collective purpose of cohort studies.

This survey provides a snapshot of how cohort researchers perceive and facilitate impact across diverse projects and settings, to inform further inquiry on this priority issue for the field.

The breadth of contributions reported by participating researchers reflects the omnibus design of many cohort studies, which are intended to address diverse and evolving societal needs.^25, 28–30^ Consistent with anecdotal reports,^7, 17, 31^ the strongest perceived contributions were in the research domain, highlighting the central role of cohorts in advancing methodological innovation in areas like measurement development.^32^ Policy was the most frequently cited domain of contribution beyond academia, reflecting the long-standing influence of cohort evidence on policy decision-making.^8^ Reported contributions most commonly related to health and wellbeing, followed by social equity, family relationships, and education; patterns that align closely with bibliometric analyses of cohort publications.^17^

Participating researchers reported substantial time investments in activities designed to foster contributions across these domains, reinforcing the need to understand how (not just if) impact unfolds.^10, 15, 27, 33^ On average, respondents indicated spending approximately one quarter of their working week on activities such as strategic planning, digital and social media engagement, and the development of policy–oriented outputs. The prominence of social media use and tailored communication outputs mirrors trends observed in other research fields.^16^ While such activities are widely recognised as critical to achieving impact,^11, 12, 21^ this study provides the first quantitative evidence of the scale of this typically invisible work among cohort researchers.

The findings also reinforce longstanding concerns regarding the challenges of achieving and demonstrating impact.^13, 14, 34^ Participating researchers described cohort impact as cumulative and long–term. This aligns with “contribution” models, in which impact is understood as influence within complex systems, accruing incrementally, and only partially attributable to any single study or researcher.^10, 15^ However, it sits in tension with prevailing impact assessment frameworks, which tend to privilege short–term, discrete effects that are more readily attributable to a researcher or study.^6^ Such misalignment has been argued to create pressure to overstate impact claims,^13, 14^ a concern reported by the majority of participating researchers in this study.

## Strengths and limitations

To our knowledge, this study is the first quantitative investigation of how cohort researchers understand and facilitate research impact. Data from 163 respondents, representing a cumulative 2,061 years of cohort research experience across a range of career stages and role types, provide valuable insight. While estimates should be interpreted as exploratory signals rather than precise benchmarks, this offers empirical evidence of broad patterns previously described primarily through case studies. By generating data in a field that lacks established impact frameworks, the study provides an important foundation for future methodological and empirical development.

Nevertheless, limitations warrant consideration, particularly with respect to sample selection. No single body connects all cohort researchers, necessitating a snowball sampling approach that may have introduced selection bias by favouring more networked researchers within targeted regions. Researchers more actively engaged with impact may also have been more inclined to participate, potentially upwardly biasing estimates of impact–related activity. Representativeness cannot be assessed empirically because, to our knowledge, no comprehensive data exist on the international cohort research workforce. Although the survey was internationally accessible, respondents were predominantly from Australia and the UK, likely reflecting recruitment pathways and the salience of impact policies in these settings.^34^ A larger sample would have enabled more detailed subgroup analyses across regions, career stages, and disciplines.

Objectivity is another consideration. Members of the author team are cohort researchers studying their own field, making full detachment challenging. We sought to mitigate this through collaboration with an impact researcher external to cohort studies (Author KK) and the use of pre–planned, transparent methods. Nonetheless, interpretation should consider this embeddedness. Participants similarly have a vested interest in the topic. While cohort researchers are uniquely positioned to report on their own impact–related activities and experiences navigating impact structures, they may not always be aware of downstream impacts,^20^ and responses may be shaped by a desire to emphasise challenges or present their work favourably. Efforts to minimise these risks included careful survey framing and interpretation.

## Future directions

Advancing understanding of the nature and processes of cohort impact represents an important direction for future research. This work can build on emerging empirical evidence,^9, 13, 17^ conceptual frameworks for long-term influence within complex systems,^8, 10, 15, 21^ and the many existing examples of effective cohort impact. Progress in this area will also benefit from incorporating a broader range of stakeholder perspectives, as whether and how cohort evidence informs decisions is shaped by relationships with policymakers, practitioners, communities, and other actors within the research ecosystem.^35^ A stronger evidence base can help elucidate impact processes and practices that cohort teams can realistically implement to increase the likelihood of meaningful outcomes.^36^

This strengthened evidence base can also inform more nuanced approaches to capturing cohort impact within funding and assessment structures. This attention to research structures is essential, as the challenges identified in this study are at least partly systemic, rooted in academic promotion, publishing, and funding.^31, 34^ While individual researchers have limited capacity to effect change at this level, coordinated action can help reshape research systems. Cohort networks are well placed to address cross-cutting issues,^3, 37^ including impact, but may benefit from the development of “networks of networks” that link geographic and disciplinary hubs. This could strengthen collective influence while enabling coordinated research and methodological learning across cohorts.

## Conclusions

This study highlights the breadth of cohort contributions and the processes through which researchers seek to achieve societal impact. Cohort impact is described as cumulative, unfolding over long timescales, and rarely attributable to single studies; features that sit uneasily within short-term, attribution-focused models. Limited time and resources are seen to further complicate impact practice, and most respondents reported that pressure exists to overstate impact claims. As the first quantitative snapshot of cohort researcher views, our findings can inform debate and further inquiry on the nature of impact in this context, what frameworks and metrics can credibly capture it, and how funders and institutions can better support impact. Engaging with these questions is a priority for sustaining the cohort infrastructures that underpin life–course epidemiology.

## Data Availability

Data can be requested via the Melbourne Children's LifeCourse Initiative

https://lifecourse.melbournechildrens.com/data-access/

## Acknowledgements

Work conducted at the Murdoch Children’s Research Institute is supported by the Victorian Government’s Operational Infrastructure Support Program. CO was supported by a National Health and Medical Research Council (NHMRC) Investigator Grant (GNT1175086). DB, CBS and ET are funded by the Economic and Social Research Council (ES/W013142/1).

## Conflict of interest

The Author(s) declare(s) that there is no conflict of interest.

## Research ethics statement

The study was approved as negligible risk by the Royal Children’s Hospital Human Research Ethics Committee.

## Data availability

Data can be requested via the Melbourne Children’s LifeCourse Initiative (https://lifecourse.melbournechildrens.com/data-access/; select “other” in requested data).

## Code availability

Available at https://osf.io/j4qxd/overview?view_only=418581796e5a4492ab74dd569dbb0ad2.

## Notes

### Competing Interest Statement

The authors have declared no competing interest.

