## Supplementary File 1 for "Researcher perspectives on the value and impact of population-based cohort studies"

murdoch children's research institute

AAA

### Research impact in population-based longitudinal cohort studies

#### What's the true value of cohorts? We need your voice to evolve and tailor approaches to research impact in cohort studies

Do you work with population-based longitudinal cohort studies ("cohorts") that follow groups of individuals over periods of the life course? If so, your voice can help us understand the true value that cohorts offer society. By sharing your insights, you'll help us explore new and more meaningful ways to capture the impact of cohort studies.

**Take our short 15-minute survey to share your perspectives on research impact in cohorts.** Whether you're early in your career or a senior leader, and whether your work involves data analysis, coordination, management, or other contributions to cohort studies, your input is invaluable. Multiple responses from the same cohort team are welcome, as different perspectives only enrich the conversation. Please also feel free to forward this invitation to others in your network.

**Help advance the conversation.** We understand that the shift towards impact is creating opportunities as well as challenges for cohort researchers and teams. This is the first quantitative study designed to explore researchers' practices and experiences related to cohort impact. Findings will be relevant to the cohort research community and will be shared through peer-reviewed publications and presentations.

**This survey is completely anonymous.** No identifying information about you or the cohort(s) you work with will be collected. We'll ask only general questions about your research to ensure your identity and your study remain confidential. Participation is entirely voluntary, and you may exit the survey at any time without consequences. Whether you choose to participate will not affect your relationship with our team or the cohort collaborations we represent.

This study has been deemed negligible risk by the Royal Children's Hospital HREC. By completing the survey, you are indicating your consent to participate in this research. Anonymous data from this project may be shared with approved researchers for future analyses, following appropriate ethical and institutional review.

For any questions please contact Dr Meredith O'Connor. If you have ethical concerns, contact the Royal Children's Hospital.

**Thank you for helping shape the future of cohort research impact. We truly appreciate your time and contribution.**

---

**Over the past 12 months, which of the following best describes your involvement with cohort research?** *Tick all that apply. By "cohort", we are referring to population-based longitudinal studies that collect data about a group of individuals over time.*

* must provide value

- ☐ Senior leadership as a cohort director, chief or associate investigator, or similar
- ☐ Project coordination and operations
- ☐ Lived experience/consumer advisor
- ☐ Analysis and research using cohort data
- ☐ Other active role/s in cohort research

- [ ] No involvement in cohort research in the past 12 months

#### Your role and perspective

In the following, please tell us about general aspects of your role so that we can understand more about your perspective.

**How long have you been working in cohort research?**
*Round to the nearest whole year*

__________ year(s)

**Which of the following best describe the discipline of your cohort research?** *Tick all that apply*

- [ ] Biostatistics and data science
- [ ] Demography
- [ ] Economics
- [ ] Education
- [ ] Epidemiology and public health
- [ ] Genetics and molecular biology
- [ ] Medicine and biomedical sciences
- [ ] Psychology
- [ ] Sociology
- [ ] Other

**Do you hold any concurrent appointments outside of the research/academic sector?** *Tick all that apply*

- [ ] No, I do not hold any concurrent appointments outside the research/academic sector
- [ ] Clinical (e.g., medical doctor, psychologist, allied health professional)
- [ ] Government (e.g., policy analyst, public sector strategist)
- [ ] Corporate (e.g., consultancy, pharmaceuticals industry)
- [ ] Nonprofit (e.g., social impact initiatives, philanthropic organisation)
- [ ] Other role outside of the research / academic sector

**What country do you primarily work in?**

In the following, please tell us about the cohort (i.e., study following a group of individuals over time) that has been the greatest focus of your active role/s in cohort research in the past 12 months. If you work across multiple cohorts equally, please choose one to focus on.

**What life periods does the cohort capture?** *Tick all that apply*

- ☐ Childhood (pre-conception to 10 years)
- ☐ Adolescence (11-25 years)
- ☐ Young adulthood (26-39 years)
- ☐ Middle adulthood (40-59 years)
- ☐ Later adulthood (60-79)
- ☐ Senior adulthood (80+)

**What is the approximate sample size (number of participants recruited at baseline) being followed within this cohort study?**

- ○ 499 or fewer participants
- ○ 500-999 participants
- ○ 1000-4,999 participants
- ○ 5000-9,999 participants
- ○ 10,000-19,999 participants
- ○ More than 20,000 participants

**Where is the cohort primarily based?**

#### Research impact goals, strategies, and views

In the following section, please respond regarding your cohort research program, as you define it, to best reflect your perspective. For instance, if you are part of team managing the cohort you described previously, you might focus on the impact of that cohort as whole. Alternatively, if you are a secondary data user, you might respond in relation to your own research program using data from the cohort you described.

There are no right or wrong answers. We are interested in your unique, subjective perspectives on impact, rather than impact metrics. Cohort research is not expected to impact all areas or achieve every goal; your feedback will help to highlight areas of greater or lesser focus. Your responses are anonymous and will not be attributed to your cohort.

**To what extent does your cohort research focus on the following societal issues?** We are interested in your views on the current focus and objectives of your cohort research, rather than measurable progress in tackling these large-scale social issues.

| | Not at all | Slightly | Moderately | Strongly | Not sure |
|---|---|---|---|---|---|
| **Health and wellbeing** (e.g. mental and physical health, substance use) | ○ | ○ | ○ | ○ | ○ |
| **Family relationships** (e.g. parenting, family functioning) | ○ | ○ | ○ | ○ | ○ |

| | | | | | |
|---|---|---|---|---|---|
| **Education** (e.g. curriculum, bullying, attainment) | ○ | ○ | ○ | ○ | ○ |
| **Workplace wellbeing** (e.g. stress, career pathways) | ○ | ○ | ○ | ○ | ○ |
| **Community and social networks** (e.g. social cohesion, peer support) | ○ | ○ | ○ | ○ | ○ |
| **Crime and justice** (e.g., delinquency, victimisation, contact with criminal justice system) | ○ | ○ | ○ | ○ | ○ |
| **Economic factors** (e.g. cost of living, housing availability) | ○ | ○ | ○ | ○ | ○ |
| **Social equity** (e.g. discrimination, social disadvantage) | ○ | ○ | ○ | ○ | ○ |
| **Digital and technology** (e.g. online safety, cybersecurity) | ○ | ○ | ○ | ○ | ○ |
| **Historical impacts** (e.g. intergenerational trauma, population aging) | ○ | ○ | ○ | ○ | ○ |
| **Other** | ○ | ○ | ○ | ○ | ○ |

**To what extent does your cohort research contribute across the following domains?** We are interested in your views on the current focus and objectives of your cohort research, rather than measurable progress.

| | Not at all | Slightly | Moderately | Strongly | Not sure |
|---|---|---|---|---|---|
| **Research** (e.g., pursuit of new knowledge, developing new research methods or approaches, shaping the research agenda in your field, or training of junior researchers) | ○ | ○ | ○ | ○ | ○ |
| **Policy** (e.g., driving development of new policy, raising awareness and mobilising support for an issue among policymakers, or introducing new ideas, language, or concepts into policy discussions) | ○ | ○ | ○ | ○ | ○ |
| **Services** (e.g., change to treatment or targeting of intervention, informing data and reporting in health, education, social, or other service systems, improving the cost-effectiveness of services) | ○ | ○ | ○ | ○ | ○ |
| **Societal** (e.g., shaping public understanding and awareness of issues, influencing societal values and culture, and improving the public's ability to access and use information that benefits them) | ○ | ○ | ○ | ○ | ○ |
| **Other** | ○ | ○ | ○ | ○ | ○ |

**Thinking about the impact of your cohort research, how often do you observe the following?** We are interested in your perceptions of how impact typically unfolds, based on your experience in cohort research.

| | Never | Rarely | Sometimes | Almost always | Not sure |
|---|---|---|---|---|---|
| **The research leads to specific, tangible changes in policy or practice** | ○ | ○ | ○ | ○ | ○ |
| **It's possible to trace a clear, causal link between cohort research and its impact** | ○ | ○ | ○ | ○ | ○ |
| **Impact can be demonstrated shortly after the research is conducted** | ○ | ○ | ○ | ○ | ○ |
| **Impact arises from a single piece of research (as opposed to a body of work)** | ○ | ○ | ○ | ○ | ○ |

**What strategies does your cohort research program employ to try to achieve impact?** We do not expect all cohorts to employ all strategies, nor do we assume that these strategies necessarily translate into measurable impact.

| | Never | Rarely | Sometimes | Almost always | Not sure |
|---|---|---|---|---|---|
| **Strategic planning** (e.g., co-defining impact goals with stakeholders; planning and embedding impact evaluation) | ○ | ○ | ○ | ○ | ○ |
| **Academic dissemination** (e.g., peer-reviewed articles; conferences or symposia; segments in institutional newsletters) | ○ | ○ | ○ | ○ | ○ |
| **Policy reports** (e.g., producing summaries of evidence in relation to policy, contributing submissions to government inquiries) | ○ | ○ | ○ | ○ | ○ |
| **Community forums** (e.g., accessible talks, workshops or training sessions for community groups) | ○ | ○ | ○ | ○ | ○ |
| **Traditional media** (e.g., sharing evidence through broadcast or print via major news outlets) | ○ | ○ | ○ | ○ | ○ |
| **Digital and social media** (e.g., podcasts, LinkedIn posts, Twitter/X threads, short-form videos) | ○ | ○ | ○ | ○ | ○ |
| **Cross-sector partnerships** (e.g., undertaking research with industry or NGO partners) | ○ | ○ | ○ | ○ | ○ |

| | | | | | |
|---|---|---|---|---|---|
| **Parliamentary engagement** (e.g., roundtables that connect researchers and policymakers) | ○ | ○ | ○ | ○ | ○ |
| **Other** | ○ | ○ | ○ | ○ | ○ |

**What time investment does impact require?** The activities above can involve time intensive tasks such as meetings or emails with stakeholders, development of translational outputs, and strategising about impact.

**In a typical week, what proportion of your total work time do you spend on these activities?**

0% 50% 100%

Change the slider above to set a response

reset

**In an ideal week, what percentage of your work time would you like to spend on these activities?**

0% 50% 100%

Change the slider above to set a response

reset

**To what extent do you agree with the following statements about how research impact is currently approached?** We are interested in your views on the current research environment, which includes research policy, funders, journals, and research organisations.

| | Strongly disagree | Somewhat disagree | Somewhat agree | Strongly agree | Not sure |
|---|---|---|---|---|---|
| **Current impact approaches align well with the goals and motivations of cohort research** | ○ | ○ | ○ | ○ | ○ |
| **Existing impact frameworks capture and value the types of societal impact that cohort studies typically generate** | ○ | ○ | ○ | ○ | ○ |
| **Pressure exists for cohort researchers to overstate claims about impact** | ○ | ○ | ○ | ○ | ○ |
| **Cohort studies are disadvantaged compared to other study designs by current impact evaluation approaches** | ○ | ○ | ○ | ○ | ○ |
| **Skills, resources, and support are available to cohort teams to plan, enable, and monitor impact** | ○ | ○ | ○ | ○ | ○ |

**Any further reflections**

**Would you like to offer any other reflections regarding impact in the context of cohort studies?**
For example, we are interested to hear about other barriers you face, whether you feel adequately resourced in this area, how you are approaching specific areas like monitoring your impact, or specific examples of impact that you may wish to share.

#### Thank you for your participation

We appreciate the time you have taken and thank you for being part of this study.

Our goal is to provide insights on the nature of research impact in the context of cohort studies.

We can't achieve this without the essential perspectives of you and others involved in cohort research, who know best the impact intentions and challenges of your work. Please also feel free to distribute this survey link to your colleagues and collaborators.

The results will be disseminated through academic publications, conference presentations, and via the Melbourne Children's LifeCourse Initiative.

If you have any questions, feel free to contact the lead researcher Dr Meredith O'Connor.

Submit

Powered by REDCap - Cookie policy
