## Supplementary File 2 for "Researcher perspectives on the value and impact of population-based cohort studies"

Supplementary Table 1. Missing data on study variables.

| Variable | N (%) with missing data |
|---|---|
| **Researcher characteristics** | |
| Role type | 0 (0%) |
| Years in cohort research | 0 (0%) |
| Primary country in which working | 0 (0%) |
| Field of Research | 0 (0%) |
| Concurrent appointments | 0 (0%) |
| **Cohort the researcher primarily works with** | |
| Life periods captured | 0 (0%) |
| Sample size of cohort at recruitment | 1 (0.6%) |
| Primary country in which cohort is based | 4 (2.5%) |
| **To what extent does your cohort research contribute to the following domains?** | |
| Research (e.g., pursuit of new knowledge, developing new research methods or approaches, shaping the research agenda in your field, or training of junior researchers) | 2 (1.2%) |
| Policy (e.g., driving development of new policy, raising awareness and mobilising support for an issue among policymakers, or introducing new ideas, language, or concepts into policy discussions) | 1 (0.6%) |
| Services (e.g., change to treatment or targeting of intervention, informing data and reporting in health, education, social, or other service systems, improving the cost-effectiveness of services) | 5 (3.1%) |
| Societal (e.g., shaping public understanding and awareness of issues, influencing societal values and culture, and improving the public's ability to access and use information that benefits them) | 5 (3.1%) |
| **To what extent does your cohort research focus on the following societal issues?** | |
| Health and wellbeing (e.g. mental and physical health, substance use) | 2 (1.2%) |
| Family relationships (e.g. parenting, family functioning) | 3 (1.8%) |
| Education (e.g. curriculum, bullying, attainment) | 1 (0.6%) |
| Workplace wellbeing (e.g. stress, career pathways) | 8 (4.9%) |
| Community and social networks (e.g. social cohesion, peer support) | 5 (3.1%) |
| Crime and justice (e.g., delinquency, victimisation, contact with criminal justice system) | 8 (4.9%) |
| Economic factors (e.g. cost of living, housing availability) | 5 (3.1%) |
| Social equity (e.g. discrimination, social disadvantage) | 5 (3.1%) |
| Digital and technology (e.g. online safety, cybersecurity) | 8 (4.9%) |
| Historical impacts (e.g. intergenerational trauma, population aging) | 9 (5.5%) |
| **Thinking about the impact of your research, how often do you observe the following?** | |
| The research leads to specific, tangible changes in policy or practice | 5 (3.1%) |
| It's possible to trace a clear, causal link between cohort research and its impact | 5 (3.1%) |
| Impact can be demonstrated shortly after the research is conducted | 7 (4.3%) |
| Impact arises from a single piece of research (as opposed to a body of work) | 8 (4.9%) |
| **What strategies does your cohort research program employ to try to achieve impact?** | |
| Strategic planning (e.g., co-defining impact goals with stakeholders; planning and embedding impact evaluation) | 6 (3.7%) |
| Academic dissemination (e.g., peer-reviewed articles; conferences or symposia; segments in institutional newsletters) | 1 (0.6%) |
| Policy reports (e.g., producing summaries of evidence in relation to policy, contributing submissions to government inquiries) | 6 (3.7%) |
| Community forums (e.g., accessible talks, workshops or training sessions for community groups) | 9 (5.5%) |
| Traditional media (e.g., sharing evidence through broadcast or print via major news outlets) | 4 (2.5%) |
| Digital and social media (e.g., podcasts, LinkedIn posts, Twitter/X threads, short-form videos) | 4 (2.5%) |
| Cross-sector partnerships (e.g., undertaking research with industry or NGO partners) | 7 (4.3%) |
| Parliamentary engagement (e.g., roundtables that connect researchers and policymakers) | 11 (6.7%) |
| **Time commitment** | |
| In a typical week, what proportion of your total work time do you spend on these activities? | 6 (3.7%) |
| In an ideal week, what percentage of your work time would you like to spend on these activities? | 7 (4.3%) |
| **To what extent do you agree with the following statements about how research impact is currently approached?** | |
| Current impact approaches align well with the goals and motivations of cohort research | 25 (15.3%) |
| Existing impact frameworks capture and value the types of societal impact that cohort studies typically generate | 25 (15.3%) |

| | |
|---|---|
| Pressure exists for cohort researchers to overstate claims about impact | 14 (8.6%) |
| Cohort studies are disadvantaged compared to other study designs by current impact evaluation approaches | 23 (14.1%) |
| Skills, resources, and support are available to cohort teams to plan, enable, and monitor impact | 19 (11.7%) |

Supplementary Table 2. Extent to which cohort research contributes to different domains and specific societal issues reported by cohort researchers based on their subjective views of the current focus and objectives of their cohort research.

| | Not at all | | | Slightly | | | Moderately | | | Strongly | | |
|---|---|---|---|---|---|---|---|---|---|---|---|---|
| | % | 95% CI | | % | 95% CI | | % | 95% CI | | % | 95% CI | |
| **To what extent does your cohort research contribute to the following domains?** | | | | | | | | | | | | |
| Research (e.g., pursuit of new knowledge, developing new research methods or approaches, shaping the research agenda in your field, or training of junior researchers) | 0.00 | 0.00 | 0.00 | 4.97 | 2.49 | 9.67 | 11.18 | 7.13 | 17.10 | 83.85 | 77.28 | 88.80 |
| Policy (e.g., driving development of new policy, raising awareness and mobilising support for an issue among policymakers, or introducing new ideas, language, or concepts into policy discussions) | 3.09 | 1.28 | 7.24 | 19.75 | 14.29 | 26.66 | 46.30 | 38.71 | 54.06 | 30.86 | 24.19 | 38.45 |
| General public (e.g., shaping public understanding and awareness of issues, influencing societal values and culture, and improving the public's ability to access and use information that benefits them) | 3.80 | 1.71 | 8.24 | 29.11 | 22.52 | 36.73 | 40.51 | 33.08 | 48.39 | 26.58 | 20.24 | 34.07 |
| Services (e.g., change to treatment or targeting of intervention, informing data and reporting in health, education, social, or other service systems, improving the cost-effectiveness of services) | 12.03 | 7.78 | 18.14 | 33.54 | 26.57 | 41.32 | 35.44 | 28.33 | 43.26 | 18.99 | 13.57 | 25.92 |
| **To what extent does your cohort research focus on the following societal issues?** | | | | | | | | | | | | |
| Health and wellbeing (e.g. mental and physical health, substance use) | 1.86 | 0.60 | 5.66 | 2.48 | 0.93 | 6.48 | 7.45 | 4.26 | 12.71 | 88.20 | 82.19 | 92.37 |
| Family relationships (e.g. parenting, family functioning) | 14.37 | 9.71 | 20.76 | 23.13 | 17.20 | 30.34 | 30.63 | 23.93 | 38.25 | 31.87 | 25.08 | 39.55 |
| Social equity (e.g. discrimination, social disadvantage) | 12.66 | 8.29 | 18.86 | 30.38 | 23.67 | 38.05 | 26.58 | 20.24 | 34.07 | 30.38 | 23.67 | 38.05 |
| Education (e.g. curriculum, bullying, attainment) | 19.14 | 13.76 | 25.98 | 27.16 | 20.83 | 34.58 | 25.31 | 19.17 | 32.62 | 28.40 | 21.94 | 35.87 |
| Economic factors (e.g. cost of living, housing availability) | 17.09 | 11.95 | 23.83 | 26.58 | 20.24 | 34.07 | 30.38 | 23.67 | 38.05 | 25.95 | 19.67 | 33.40 |
| Community and social networks (e.g. social cohesion, peer support) | 20.25 | 14.66 | 27.30 | 33.54 | 26.57 | 41.32 | 31.65 | 24.82 | 39.36 | 14.56 | 9.84 | 21.01 |
| Historical impacts (e.g. intergenerational trauma, population aging) | 42.21 | 34.60 | 50.20 | 22.73 | 16.75 | 30.07 | 20.78 | 15.05 | 27.97 | 14.29 | 9.56 | 20.80 |
| Workplace wellbeing (e.g. stress, career pathways) | 45.16 | 37.45 | 53.11 | 25.16 | 18.91 | 32.64 | 20.00 | 14.39 | 27.10 | 9.68 | 5.90 | 15.49 |
| Digital and technology (e.g. online safety, cybersecurity) | 59.35 | 51.39 | 66.86 | 23.23 | 17.20 | 30.58 | 10.97 | 6.90 | 16.99 | 6.45 | 3.49 | 11.63 |
| Crime and justice (e.g., delinquency, victimisation, contact with criminal justice system) | 64.52 | 56.62 | 71.70 | 22.58 | 16.64 | 29.89 | 9.03 | 5.40 | 14.73 | 3.87 | 1.74 | 8.40 |

Supplementary Table 3. Processes to achieve impact reported by cohort researchers, based on their subjective viewpoint. Does not assume that these strategies are successful in achieving impact.

| | Never | | | Rarely | | | Sometimes | | | Almost always | | |
|---|---|---|---|---|---|---|---|---|---|---|---|---|
| | % | 95% CI | | % | 95% CI | | % | 95% CI | | % | 95% CI | |
| **Thinking about the impact of your research, how often do you observe the following?** | | | | | | | | | | | | |
| The research leads to specific, tangible changes in policy or practice | 0.63 | 0.09 | 4.42 | 26.58 | 20.24 | 34.07 | 65.19 | 57.38 | 72.26 | 7.59 | 4.34 | 12.95 |
| It's possible to trace a clear, causal link between cohort research and its impact | 2.53 | 0.95 | 6.60 | 32.28 | 25.41 | 40.01 | 55.70 | 47.81 | 63.30 | 9.49 | 5.78 | 15.20 |
| Impact can be demonstrated shortly after the research is conducted | 8.97 | 5.37 | 14.64 | 58.33 | 50.39 | 65.86 | 30.13 | 23.40 | 37.84 | 2.56 | 0.96 | 6.68 |
| Impact arises from a single piece of research (as opposed to a body of work) | 17.42 | 12.19 | 24.27 | 49.68 | 41.82 | 57.55 | 28.39 | 21.80 | 36.04 | 4.52 | 2.16 | 9.22 |
| **What strategies does your cohort research program employ to try to achieve impact?** | | | | | | | | | | | | |
| Academic dissemination (e.g., peer-reviewed articles; conferences or symposia; segments in institutional newsletters) | 0.00 | 0.00 | 0.00 | 1.85 | 0.59 | 5.63 | 9.26 | 5.64 | 14.84 | 88.89 | 83.00 | 92.91 |
| Strategic planning (e.g., co-defining impact goals with stakeholders; planning and embedding impact evaluation) | 8.28 | 4.85 | 13.79 | 14.01 | 9.38 | 20.42 | 43.95 | 36.33 | 51.86 | 33.76 | 26.75 | 41.56 |
| Digital and social media (e.g., podcasts, LinkedIn posts, Twitter/X threads, short-form videos) | 6.29 | 3.40 | 11.34 | 12.58 | 8.23 | 18.75 | 48.43 | 40.70 | 56.23 | 32.70 | 25.82 | 40.43 |
| Traditional media (e.g., sharing evidence through broadcast or print via major news outlets) | 4.40 | 2.10 | 8.99 | 18.87 | 13.48 | 25.76 | 55.35 | 47.49 | 62.94 | 21.38 | 15.66 | 28.50 |
| Policy reports (e.g., producing summaries of evidence in relation to policy, contributing submissions to government inquiries) | 4.46 | 2.13 | 9.10 | 21.66 | 15.86 | 28.84 | 52.87 | 44.99 | 60.60 | 21.02 | 15.31 | 28.15 |
| Community forums (e.g., accessible talks, workshops or training sessions for community groups) | 12.34 | 7.98 | 18.59 | 31.17 | 24.31 | 38.97 | 44.16 | 36.46 | 52.14 | 12.34 | 7.98 | 18.59 |
| Cross-sector partnerships (e.g., undertaking research with industry or NGO partners) | 16.67 | 11.57 | 23.41 | 26.92 | 20.50 | 34.48 | 45.51 | 37.81 | 53.44 | 10.90 | 6.86 | 16.89 |
| Parliamentary engagement (e.g., roundtables that connect researchers and policymakers) | 19.74 | 14.12 | 26.89 | 41.45 | 33.83 | 49.50 | 33.55 | 26.45 | 41.49 | 5.26 | 2.64 | 10.22 |

Supplementary Figure 1. Researchers reports on the proportion of a typical work week spent on impact related activities, according to both their actual and ideal work practices.

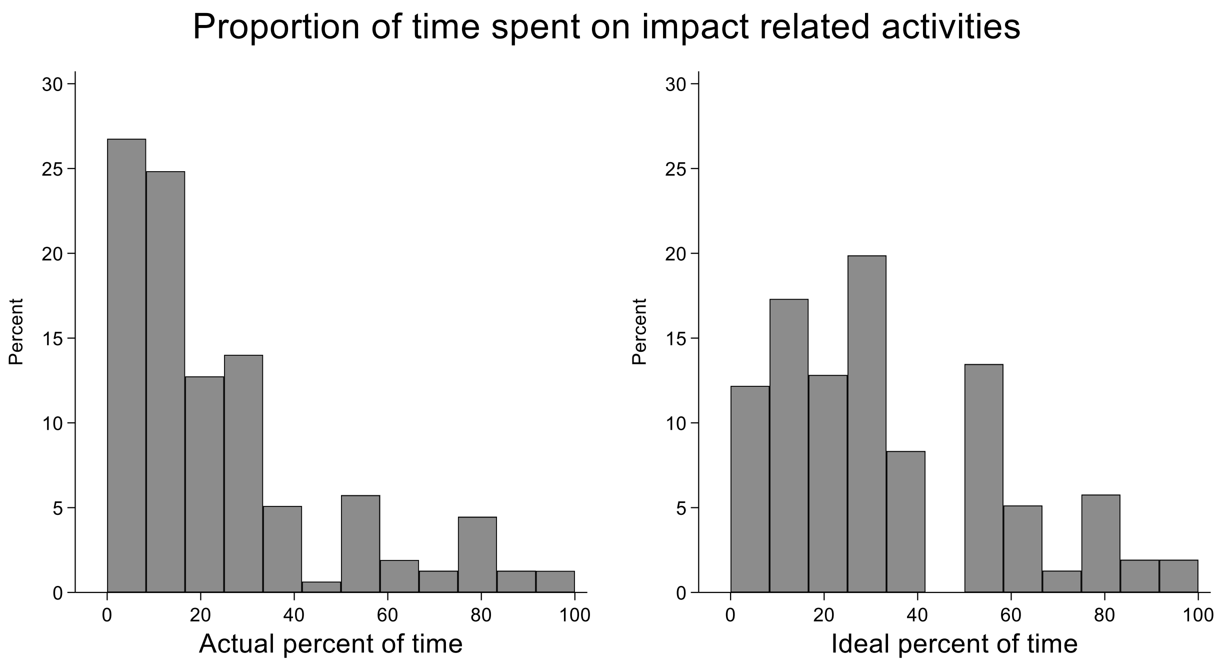


Supplementary Table 4. Challenges and opportunities in impact as reported by cohort researchers.

| To what extent do you agree with the following statements about how research impact is currently approached? | Strongly disagree | | | Somewhat disagree | | | Somewhat agree | | | Strongly agree | | |
|---|---|---|---|---|---|---|---|---|---|---|---|---|
| | % | 95% CI | | % | 95% CI | | % | 95% CI | | % | 95% CI | |
| Current impact approaches align well with the goals and motivations of cohort research | 4.35 | 1.95 | 9.40 | 36.96 | 29.26 | 45.38 | 46.38 | 38.16 | 54.79 | 12.32 | 7.76 | 18.99 |
| Existing impact frameworks capture and value the types of societal impact that cohort studies typically generate | 13.04 | 8.34 | 19.82 | 49.28 | 40.96 | 57.63 | 31.88 | 24.60 | 40.18 | 5.80 | 2.91 | 11.23 |
| Pressure exists for cohort researchers to overstate claims about impact | 2.68 | 1.00 | 6.99 | 17.45 | 12.13 | 24.46 | 50.34 | 42.30 | 58.35 | 29.53 | 22.71 | 37.40 |
| Cohort studies are disadvantaged compared to other study designs by current impact evaluation approaches | 3.57 | 1.48 | 8.35 | 18.57 | 12.92 | 25.95 | 50.00 | 41.72 | 58.28 | 27.86 | 21.01 | 35.92 |
| Skills, resources, and support are available to cohort teams to plan, enable, and monitor impact | 27.78 | 21.03 | 35.72 | 36.81 | 29.27 | 45.04 | 26.39 | 19.79 | 34.25 | 9.03 | 5.29 | 14.99 |
